# Use of large language models by academic hospitalists: results of a multicenter survey

**DOI:** 10.64898/2026.05.27.26353610

**Authors:** Eric Bressman, Andrew D. Auerbach, Angela Keniston, Caroline Jens, Sumant R. Ranji

## Abstract

**Introduction:** The use of artificial intelligence (AI) by clinicians has increased rapidly in recent years, with large language models (LLMs) emerging as tools that can equal clinician diagnostic performance in simulated settings. However, limited data exist regarding physicians use of LLMs in real-world clinical practice. This study aimed to evaluate the frequency of LLM use among practicing hospitalists, identify which LLMs are most commonly utilized, and assess hospitalists’ perceptions of the benefits and limitations of LLM use in clinical care.

**Methods:** We conducted a cross-sectional survey study of academic hospital medicine faculty across 8 institutions within the Hospital Medicine Reengineering Network (HOMERuN), a collaborative research consortium. Eligible participants included hospitalists practicing within participating HOMERuN sites during the study period. The survey assessed the frequency of LLM use, types of LLMs used, clinical applications, and physician perceptions regarding usefulness, efficiency, and concerns associated with LLM adoption.

**Results:** 170 respondents (67.1%) reported ever using an LLM in clinical practice. Among LLM users, OpenEvidence was the most used tool (88.9%), followed by ChatGPT (58.5%), Google Gemini (26.9%), and Microsoft Copilot (20.5%). Only a minority of hospitalists reported using LLMs daily while seeing patients. The most common use cases of LLMs were answering diagnostic (77.1%) and management (77.6%) questions. A majority also reported using LLMs to identify or summarize primary literature (60.0%). Lack of trust in outputs (49.8%), uncertainty around institutional policies (48.6%), and lack of access to secure applications(43.1%) were cited as the most frequent barriers to using LLMs in practice.

**Discussion:** The use of LLMs in clinical practice is already widespread, though regular or daily use is not yet typical. Concerns regarding reliability, patient privacy, and safe integration into clinical workflows remain significant barriers to broader adoption. The responsible implementation of LLMs in hospital medicine will require addressing these barriers.

## Introduction

The use of artificial intelligence (AI) by clinicians has rapidly increased in the past 2 years. According to recent survey data from the American Medical Association,^1^ over 80% of physicians currently use some form of AI in their practice for an expanding range of use cases. Concurrently, advances in generative AI - particularly large language models (LLMs)-have yielded applications that can perform complex clinical activities, including diagnostic and management reasoning for hospitalized patients. Multiple studies^2,3^ in simulated settings show that LLMs can equal practicing clinician’s performance on these tasks. Clinicians now have access to a range of LLMs and LLM-based tools, and at many institutions, secure LLMs are approved for use with protected health information (PHI) and being integrated into electronic health records.

Though clinicians are increasingly using AI, few studies have investigated how physicians are currently using large language models at the point of care or how practicing clinicians perceive the benefits and drawbacks of using LLMs.^4–6^ There are over 50,000 hospitalists in the United States,^7^ making the field one of the 5 largest medical specialties. In this study, we surveyed academic hospitalists regarding their use of LLMs in their clinical practice. The goals of the survey were to establish the prevalence and frequency of LLM usage among practicing hospitalists; identify which LLMs hospitalists use and for what tasks; obtain their perceptions of barriers to integrating LLMs into their daily work; and understand hospitalists’ perceptions of the potential benefits and potential harms of LLM usage in clinical care.

## Methods

### Study Design

We conducted a cross-sectional survey study of academic hospital medicine faculty across eight institutions within the Hospital Medicine Reengineering Network^8^ (HOMERuN), a collaborative research consortium. The goal was to characterize patterns of large language model (LLM) use in clinical care, perceived utility of LLMs relative to standard clinical resources, and barriers to LLM adoption among hospitalists.

The protocol was reviewed by the Institutional Review Board at the University of California San Francisco and deemed exempt. We report the study results according to the CROSS guidelines.^9^

### Participants and Eligibility

Eligible participants were physician hospitalists employed within participating HOMERuN sites during the survey window. We defined hospitalists as attending physicians whose primary faculty appointment was in the Division or Section of Hospital Medicine at each participating institution. Advanced practice providers and trainees were not targeted in the distribution lists; responses indicating non-attending physician roles were excluded from analyses a priori.

### Survey Development

We developed a novel survey instrument designed to assess hospitalists’ experiences with and perceptions of LLMs in clinical practice (see Appendix for text of full survey). Survey domains included:

1. Access to and availability of institutionally supported, protected health information (PHI)-approved LLM platforms
2. Typical frequency of LLM use during clinical service
3. Perceived utility of LLMs relative to established clinical resources
4. Specific clinical use-cases (e.g., differential diagnosis, management questions, literature sourcing, documentation/EHR summarization, patient communication/education, and teaching materials)
5. Barriers to use (e.g., trust/accuracy, currency of information, time/effort, usability, explainability, policy clarity, and lack of secure access)
6. Limited demographics (institution, years since completion of residency, formal Clinical Informatics training, and gender identity)

The survey incorporated both multiple-choice and open-ended items. Item formats included single-select, multiple-select, Likert-type frequency scales (Never/Rarely/Sometimes/Often), and a 3–4 category comparative utility item. Two open-ended prompts elicited specific examples of benefits and harms experienced when using LLMs in clinical contexts. We included questions about LLMs that were publicly available in October 2025, when the survey was finalized.

Draft items were iteratively refined for clarity and content validity by the investigator team. Minor wording changes were made to reduce ambiguity and streamline completion time. The final instrument contained 16 items (including skip logic for non-users) and two free-text questions.

### Survey Distribution and Administration

The final survey was administered electronically using the REDCap survey platform. Eligible participants included all faculty hospitalists affiliated with the eight participating HOMERUN institutions. Individual email addresses were collected for eligible participants at sites where hospital policy allowed, in order to precisely track response rates. When this was not possible per local policy, invitations were sent to site leads for internal distribution; response rates in this case were calculated as the number of responses divided by the number of invitations sent.

Respondents were informed that participation was voluntary and anonymous, and no identifiable personal information was collected. The email accompanying the survey described its purpose as assessing how hospitalists use LLMs in clinical care and measuring perceptions of accuracy, usefulness, and potential risks. As an incentive for participation, respondents were entered into a lottery for a $200 gift card. One gift card was distributed per site. The study period spanned November - December 2025, during which faculty received an electronic survey invitation and 3 emailed reminders over 30 days.

### Analysis

Quantitative survey items were analyzed using descriptive statistics. We calculated proportions to summarize responses to survey items (e.g. prevalence, frequency of use; perceived utility of tools). In addition, we tabulated frequency of use by years-since-residency groups (at 5-year intervals since completing training) and reported row percentages. We compared distributions using χ^2^ tests. Analyses were conducted in Stata/IC 16.1.

## Results

The survey was distributed to 756 recipients at 8 academic hospitals; 255 completed the survey (response rate 33.7%; range 25.1% - 45.7% by site). Respondent characteristics are summarized in **Table 1**. Respondents were nearly evenly split by gender and averaged 9.3 (± 7.6) years since completion of residency. Few (6.7%) had formal training in clinical informatics.

**Table 1.** Respondent characteristics.

|  |  | **Overall** | **Frequent Users** | **Infrequent or non-users** |
| --- | --- | --- | --- | --- |
| **Respondents (n)** |  | 255 | 84 | 171 |
| **Years in practice, mean (SD)** |  | 9.5 (7.6) | 9.4 (7.7) | 9.5 (7.5) |
| **Years in practice, n (%)** | 0–5 years | 97 (38.0%) | 32 (36.7%) | 65 (38.0%) |
|  | 6–10 years | 68 (26.7%) | 23 (29.1%) | 45 (26.3%) |
|  | 11–15 years | 32 (12.5%) | 10 (12.7%) | 22 (12.9%) |
|  | ≥16 years | 50 (19.6%) | 17 (21.5%) | 33 (19.3%) |
|  | Unknown | 8 (3.1%) | 2 (2.4%) | 6 (3.5%) |
| **Formal training in informatics, n (%)** |  | 17 (6.7%) | 9 (10.7%) | 8 (4.7%) |
| **Gender, n (%)** | Male | 125 (49.0%) | 45 (53.6%) | 80 (46.7%) |
|  | Female | 117 (45.9%) | 34 (40.5%) | 83 (48.5%) |
|  | Prefer not to say | 13 (5.1%) | 5 (6.0%) | 8 (4.7%) |

### LLM Access and Model Use

Overall, 170 respondents (67.1%) reported ever using an LLM in clinical practice. Among LLM users, OpenEvidence was the most used tool (88.9%), followed by ChatGPT (58.5%), Google Gemini (26.9%) and Microsoft Copilot (20.5%). Among all respondents, 107 (42.0%) reported having access to an institutional LLM approved for use with protected health information (PHI), among whom 55 (53.2%) reported having used the tool in practice.

Though most hospitalists reported using an LLM at least once, fewer reported regular use, defined as “often” using a model for any purpose. (**Table 2**). Among LLM users, 50.0% reported using an LLM regularly. OpenEvidence was the tool most frequently reported as used “often” (35.1%).

**Table 2.**
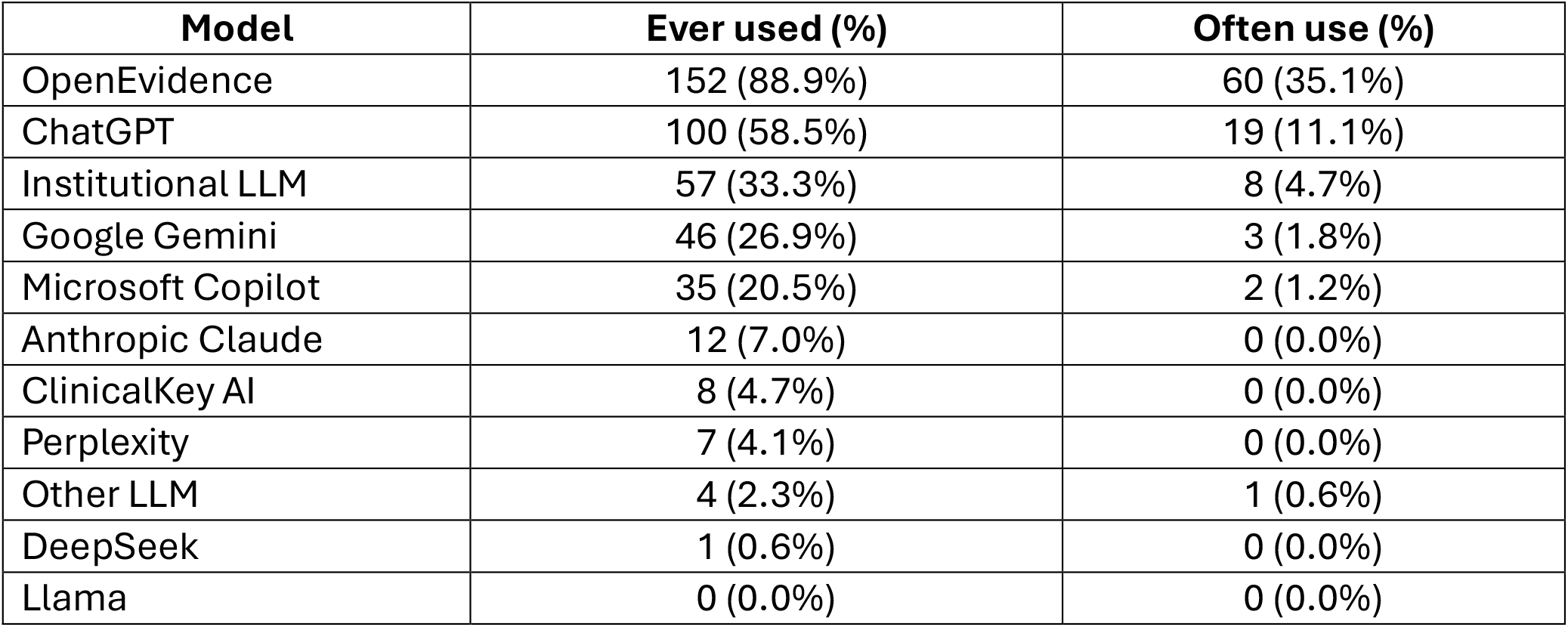
Use of Specific LLM Models Among Ever Users (n = 171)

We further characterized frequency of LLM use by asking respondents how often they used their preferred LLMs during clinical shifts **(Table 1)**. Among all LLM users, 84 (49.4%) were frequent users, defined as using an LLM at least once per clinical shift. There was no statistically significant difference in gender distribution among frequent users (53.6% of regular users identified as male and 40.5% as female, χ^2^ = 1.51, p=.471). Frequent users were distributed across all levels of clinical experience, with no difference between early- or late-career clinicians (χ^2^ = 0.32, p = .99).

### Use Cases

The most common uses of LLMs were answering diagnostic (77.1%) and management (77.6%) questions. A majority also reported using LLMs to identify or summarize primary literature (60.0%). In contrast, use for patient-facing communication (11.8%) and documentation-related tasks (≤20%) was relatively uncommon. Qualitative responses cited clinical reasoning support as a main benefit of using LLMs:

> *“I had a patient with a confusing mix of symptoms that wasn’t improving with usual treatment and basically asked [LLM] ‘what are we missing?’ A few more ideas came up that we hadn’t evaluated yet*.*”*
>
> *“A patient with moderate hyponatremia who was getting worse despite what I thought was appropriate management. The LLM helped me with my clinical assessment on why the hyponatremia management trends were proceeding as they were and helped me think through the physiology. I was able to help the patient improve with guidance from the LLM*.*”*

### Barriers to Use

Hospitalists cited multiple barriers to their use of LLMs in practice (**Table 3)**. The most frequently cited barrier was lack of trust in LLM-generated outputs (n=127, 49.8%), followed by uncertainty regarding institutional policies for LLM use (n=124, 48.6%), and lack of access to PHI-compliant tools (n=110, 43.1%). Qualitative responses highlighted concerns regarding the reliability and accuracy of LLM recommendations:

**Table 3.** Barriers to More Frequent LLM Use with Illustrative Quotes (N = 255)

| Barrier | n (%) | Illustrative Quote |
| --- | --- | --- |
| Lack of access to a PHI-approved LLM | 110 (43.1%) |  |
| Uncertainty about institutional policy | 124 (48.6%) |  |
| Institution discourages LLM use | 20 (7.8%) |  |
| Don't know how to query an LLM effectively | 92 (36.1%) | "The LLM would be most helpful in summarizing EHR information, but it is impossible to do that in a chatbot interface." |
| Don't trust the information provided by the LLM | 127 (49.8%) | "I asked [LLM] to perform a diagnostic crosscheck on a patient I was sending to MICU for acute respiratory failure. It stated that the number one differential diagnosis for the patient's respiratory failure was 'hypokalemia.' This was obviously incorrect." |
| Information provided by the LLM is not current | 49 (19.2%) | "LLM provided interesting choices for aspiration pneumonia based on outdated evidence." |
| It takes too long to input information into the LLM | 58 (22.7%) | "Takes time to generate a good query. Time to requery if first answer was not what you wanted. Time to meticulously inspect output for accuracy/errors/hallucinations. All of that time adds up." |
| Don't understand how the LLM generates its answers | 93 (36.5%) | "I have examined the output of LLM's summarizing my own clinical notes; it is clear that they do not truly understand medicine." |

**Note:**
- Respondents could select multiple barriers.

> *“My learners used [LLM] to ask a question about anticoagulation and it provided incorrect information because the LLM did not understand the specific context of the patient’s clinical needs. I had to explain that while the information provided by [the LLM] was not exactly wrong, it was wrong for our particular patient case*.*”*
>
> *“One of the downsides of LLMs*…*is that I have found I can convince it that my clinical decision making is correct, even if my clinical decision contradicts its initial advice. For example, I was trying to decide whether a patient required temporal artery biopsy for a HA (headache) that was not classic for GCA (giant cell arteritis), though the patient also had a markedly elevated ESR (erythrocyte sedimentation rate). [The LLM] recommended proceeding with temporal artery biopsy, but my gestalt was that was overly aggressive. I was able to ‘convince’ [the LLM] of this and it ultimately supported my decision to defer biopsy*.*”*

### Discussion

In this multi-center survey of academic hospitalists, we found that use of LLMs in clinical practice is already widespread, with over two-thirds of respondents reporting prior use. However, regular use remains less common, and substantial barriers to adoption persist. These findings suggest that while LLMs have rapidly entered clinical workflows, their integration into routine practice remains incomplete and uneven.

Despite widespread exposure to LLMs, only half of users reported daily use. This gap may be explained by the barriers identified in this study. Concerns about trust and accuracy were the most cited limitations, consistent with prior literature describing hallucinations and variability in LLM outputs.^10,11^ Respondents reported examples of LLMs providing advice that, if followed, could have resulted in patient harm. In addition, nearly half of respondents reported uncertainty regarding institutional policies, underscoring the importance of clearer governance frameworks.^12^ Limited access to PHI-compliant tools further constrains use in real-world clinical contexts, where data privacy considerations are paramount.

Academic hospitalists used LLMs primarily for diagnostic and management questions. Clinicians have long used digital resources (e.g., UpToDate) for these purposes, and it is likely that they are approaching LLMs (particularly curated ones, such as OpenEvidence) for similar questions due to their ability to both retrieve information and aid in interpretation and reasoning. Hospitalists’ use of LLMs as a diagnostic tool is justifiable since LLM’s diagnostic accuracy in simulated hospital medicine cases appears to be equal to practicing clinicians.^3^ Free text responses indicated that hospitalists are mainly using LLMs as a “second opinion” for diagnostically challenging cases, but as LLMs continue to improve, they may be used in earlier stages of the clinical reasoning process. It is in this respect where clinicians are likely to diverge in the degree to which they feel comfortable outsourcing these core tasks.

We found limited use of LLMs for other purposes. The low reported rate of LLM use toward patient communication is likely a function of hospitalists having relatively little electronic communication with their patients. Relatively few hospitalists reported using LLMs for chart summarization, but this is almost certainly related to limited availability of EHR-embedded tools at the time of the survey; as EHR vendors roll out HIPAA-compliant tools with access to the patient record, these use cases are likely to grow.^13^

These findings have several implications for health systems, AI application developers, and policymakers. First, as clinicians are already using LLMs for a range of clinical tasks, efforts to restrict use without providing safe alternatives may be ineffective. Instead, institutions should focus on enabling safe use through vetted tools, clear policies, and education.

Second, the prominence of trust-related concerns highlights the need for improved model evaluation, explainability, and user training. Finally, the variability in use patterns suggests that future work should examine which use cases provide the greatest clinical value and how LLMs can be optimally integrated into workflow.

This study has limitations. The sample was restricted to academic hospitalists within a single research network, which may limit generalizability. Our overall response rate was low despite multiple reminders and a financial incentive. As a result, response bias is possible, as clinicians with stronger opinions about LLMs may have been more likely to participate.

As our respondents were practicing clinicians from a large specialty, their views are likely to be a reasonable sampling of current perceptions of artificial intelligence among physicians. Additionally, self-reported use may not reflect actual usage patterns. Finally, this is a rapidly changing space with new tools and model updates being introduced on a regular basis, all of which may change usage patterns. Nonetheless, this study provides an early, multi-center snapshot of how hospitalists are incorporating LLMs into clinical care.

In conclusion, LLM use among academic hospitalists is common and focused on clinical reasoning tasks, but adoption is constrained by concerns regarding trust, policy clarity, and access to secure tools. As LLM capabilities continue to evolve, addressing these barriers will be critical to realizing their potential in clinical practice.

## Supporting information

Appendix - Survey

## Data Availability

All data produced in the present study are available upon reasonable request to the authors. The survey instrument is contained in the appendix.

## Acknowledgements

We thank the following for their assistance with survey distribution: Esteban Gershanik, MD, MPH (Brigham and Women’s Hospital), Margaret Fang, MD, MPH (University of California San Francisco), Katie Raffel, MD and Marisha Burden, MD, MPH (University of Colorado Anschutz Medical Center), Sandhya Tagaram, MD (University of Massachusetts Medical School), Ryan Greysen, MD, MS (University of Pennsylvania), Eduard Vasilevskis, MD, MPH (University of Wisconsin-Madison), Chase Webber, MD (Vanderbilt University Medical Center), and Katie Brooks, MD (San Francisco General Hospital). We also thank Tiffany Lee, MPH for assistance with project management.

