## Appendix - Survey for "Use of large language models by academic hospitalists: results of a multicenter survey"

Full text of survey instrument:

A large language model (LLM) is a type of generative artificial intelligence (AI) that can understand and generate natural language, and is trained on very large data sets, enabling it to perform a wide range of tasks. Some examples include OpenAI's ChatGPT and Google Gemini.

<u>LLM access</u>

1. Does your institution have a secure generative artificial intelligence platform that is approved for use with Protected Health Information? [Y/N/not sure – if N or not sure, skip to q3]
2. [If "yes" to q1] Which LLM is your institutional platform based on? [free text]
3. Does your institution have an institutional subscription to any of the following clinical evidence sources? [check all that apply]
    a. OpenEvidence
    b. ClinicalKeyAI
    c. UpToDate
    d. Other clinical evidence source [free text]

<u>LLM usage</u>

4. Have you ever used a large language model to answer clinical questions to help care for your hospital medicine patients?
    a. Yes
    b. No [jump to question 14]
5. Which of the following large language models have you used to answer clinical questions? [options: often/sometimes/rarely/never]
    a. Anthropic Claude
    b. ChatGPT (free version with access to GPT 4.0)
    c. ChatGPT Pro (paid version with access to OpenAI o1)
    d. ClinicalKeyAI
    e. DeepSeek
    f. Google Gemini
    g. Llama
    h. Microsoft Copilot
    i. My institution's secure LLM
    j. OpenEvidence
    k. Perplexity
    l. Other LLM not listed above [free text]
6. What type of questions do you use the LLM to answer? [options: often/sometimes/rarely/never]
    a. Diagnostic questions
    b. Therapeutic questions
    c. Questions on communicating with patients
    d. Other types of questions [free text]
7. For the LLM you use most frequently, how often during a typical clinical rotation do you use the LLM?
    a. Multiple times per shift

   b. About once per shift
   c. A few times per week
   d. A few times per month
   e. Other [free text]
8. What other resources do you typically use for help with answering clinical questions? [check all that apply]
   a. UpToDate
   b. Other online textbooks [specify]
   c. Searching Google or another similar search engine
   d. Other resource [free text]
9. How would you rate the utility of a typical LLM compared to other, non-LLM clinical resources in helping you care for your patients?
   a. I find standard clinical resources to be more helpful
   b. I find LLM's to be more useful
   c. It's hard to say which one is better
10. Which of the following are barriers to your using LLM's more frequently in clinical practice? [yes/no for each option]
   a. I do not have access to a secure LLM that is approved for PHI
   b. I'm not sure what my institution's policy is on LLM use for clinical purposes
   c. My institution explicitly discourages using LLM's for clinical purposes
   d. I don't know how to query an LLM effectively
   e. I don't trust the information provided by the LLM
   f. The information provided by the LLM is not current
   g. It takes too long to input information into the LLM
   h. I don't understand how the LLM generates its answers
   i. Other [optional, free text]
11. If your institution has a secure LLM that is approved for use with PHI, which of the following are barriers to your using it more frequently in clinical practice? [yes/no for each option]
   a. I am still concerned about violating PHI by using the LLM (even though it is approved)
   b. I don't know how to query an LLM effectively
   c. I don't trust the information provided by the LLM
   d. The information provided by the LLM is not current
   e. It takes too long to input information into the LLM
   f. I don't understand how the LLM generates its answers
   g. Other [optional, free text]
   h. Does not apply
12. Optional: Please provide an example from your practice of a clinical scenario where an LLM helped you provide better care for your patient. Do not include any PHI and do not access the electronic medical record to provide information regarding the scenario. [free text]
13. Optional: Please provide an example from your practice of a clinical scenario where an LLM provided misleading or incorrect information that could have caused harm to a patient. Do not include any PHI and do not access the electronic medical record to provide information regarding the scenario. [free text]

<u>Demographic questions</u>

14. Institution [dropdown menu with participating institutions]

15. Years since completed residency training [free text]
16. Do you have formal training (course work or fellowship) in Clinical Informatics? [Y/N]
17. Have you received any training in the use of large language models? [Y/N]
18. Gender identity [M/W/NB/choose not to state]
